# Optimal Pharmacological Management and Prevention of Glucocorticoid-Induced Osteoporosis (GIOP): Protocol for a Systematic Review and Network Meta-Analysis

**DOI:** 10.1101/19010520

**Authors:** Jiawen Deng, Emma Huang, Zachary Silver, Elena Zheng, Kyra Kavanaugh, Aaron Wen, Stephanie Sanger, Johanna Dobransky, George Grammatopoulos

## Abstract

**Introduction:** Glucocorticoid (GC) administration is an effective therapy commonly used in the treatment of autoimmune and inflammatory diseases. However, the use of GC can give rise to serious complications. The main detrimental side effect of GC therapy is significant bone loss, resulting in glucocorticoid-induced osteoporosis (GIOP).

There are a variety of treatments available for preventing and managing GIOP; however, without clearly defined guidelines, it can be very difficult for physicians to choose the optimal therapy for their patients. Previous network meta-analyses (NMAs) and meta-analyses did not include all available RCT trials, or only performed pairwise comparisons. We present a protocol for a NMA that incorporates all available RCT patient data to provide the most comprehensive ranking of all available GIOP treatments in terms of their ability to increase bone mineral density (BMD) and decrease fracture incidences among adult patients undergoing GC treatments.

**Methods and Analysis:** We will search MEDLINE, EMBASE, PubMed, Web of Science, CINAHL, CENTRAL and Chinese literature sources (CNKI, CQVIP, Wanfang Data, Wanfang Med Online) for randomized controlled trials (RCTs) which fit our criteria. RCTs that evaluate different antiresorptive regimens taken by adult patients undergoing GC therapy during the study or had taken GC for at least 3 months in the year prior to study commencement with lumbar spine BMD, femoral neck BMD, total hip BMD, vertebral fracture incidences and/or non-vertebral fracture incidences as outcomes will be selected.

We will perform title/abstract and full-text screening as well as data extraction in duplicate. Risk of bias (ROB) will be evaluated in duplicate for each study, and the quality of evidence will be examined using CINeMA in accordance to the GRADE framework. We will use R and gemtc to perform the NMA. We will report BMD results as weighted mean differences (WMDs) and standardized mean differences (SMDs), and we will report fracture incidences as odds ratios. We will use the surface under the cumulative ranking curve (SUCRA) scores to provide numerical estimations of the rankings of interventions.

**Ethics and Dissemination:** The study will not require ethical approval. The findings of the NMA will be disseminated in a peer-reviewed journal and presented at conferences. We aim to produce the most comprehensive quantitative analysis regarding the management of GIOP. Our analysis should be able to provide physicians and patients with an up-to-date recommendation for pharmacotherapies in reducing incidences of bone loss and fractures associated with GIOP.

**Systematic Review Registration:** International Prospective Register for Systematic Reviews (PROSPERO) — CRD42019127073

**ARTICLE SUMMARY:** *Strengths and limitations of this study:*

- Literature search in Chinese databases will likely yield huge amounts of new RCT evidence regarding GIOP
- Reporting change in BMD outcomes as standardized mean differences allow the pooling of absolute and percentage change data, increasing the number of RCT trials included
- Only RCTs will be included, quality of trials and networks will be evaluated using Risk of Bias and GRADE
- Older trials may report inaccurate results due to outdated procedures and hardware
- Chinese clinicians may not use the same procedures and practices as Western clinicians

## INTRODUCTION

Glucocorticoid (GC) is a class of steroids commonly prescribed to patients suffering from chronic inflammatory diseases. Its immunosuppressive properties and excellent ability of inhibiting a variety of inflammatory mediators make it an ideal choice for treating autoimmune diseases, such as systemic lupus erythematosus, or chronic respiratory diseases such as asthma[1, 2]. Because of these applications, the chronic use of GC is prevalent in clinical settings. In the first national estimate of oral GC usage in the US, it was reported that over 2,000,000 patients had taken GCs from 1999-2008. The mean duration of GC use was over 1,000 days, with 28% of patients having used GCs for five or more years[3].

However, chronic use of GC is often linked to serious, debilitating side effects, including skeletal, metabolic, and cardiovascular disorders[4]. One particularly worrying aspect about the chronic use of GC is its destructive effect on bones. Glucocorticoids facilitate osteoclast differentiation and inhibit factors required for osteoblast proliferation, resulting in decreased bone formation and increased bone resorption, causing glucocorticoid-induced osteoporosis (GIOP)[5]. Osteoporotic fractures occur in 30-50% of patients placed on long-term GC therapy, and the risk of fractures increases significantly during the first 3-6 months of GC use[6-9]. Bone weaknesses can occur with daily doses as low as 2.5 mg of prednisone[10].

The current recommended first line therapy for GIOP treatment is bisphosphonates, a class of antiresorptive drugs commonly used to treat a wide variety of skeletal diseases[11, 12]. This recommendation is supported by previous meta-analyses, which examined bisphosphonates’ ability to increase anti-fracture surrogate markers such as bone mineral density (BMD). However, these analyses were often limited as they only compared bisphosphonates to placebo or calcium/vitamin D control without head-to-head comparisons to other antiresorptive therapies[13, 14]. With new drugs being approved for use with GIOP, such as teriparatide and denosumab, traditional pairwise meta-analyses are no longer capable of integrating all available RCT data[15, 16].

Network meta-analyses (NMAs) enable comparisons between multiple treatment arms at once — even in the absence of direct comparisons between arms. Only a NMA is capable of integrating all available clinical trial data for a given patient population, and because NMAs generally include more patients overall, they can derive more accurate and precise estimates of treatment effects[17]. The latest NMA regarding GIOP were limited by its inclusion criteria, such the inclusion of only double blinded RCTs. As a result, the efficacies of treatments such as ibandronate, raloxifene and denosumab were difficult to evaluate due to a lack of data[18]. We propose to conduct an updated NMA without such limitations, as well as an extended literature search in major Chinese databases. Our question is: What are the comparative effects (in terms of changes in BMD and fracture incidences) of different antiresorptive pharmacotherapies on adult patients taking GCs.

## METHODS AND ANALYSIS

We will conduct the systematic review and NMA in accordance to the Preferred Reporting Items for Systematic Reviews and Meta-Analyses (PRISMA) incorporating NMA of health care interventions[19]. This study is prospectively registered on The International Prospective Register of Systematic Reviews (PROSPERO) — CRD42019127073. Any significant amendments to this protocol will be reported and published with the results of the review.

### Eligibility Criteria

#### Types of Participants

Adult patients (18 years or older) who had taken GC therapy for at least 3 months in the year prior to study commencement, or will be taking GC therapy during the study for at least 3 months. There is no limitation on the type or route of GC therapy. Trials were excluded if they include patients with fatal diseases or organ transplant patients, but otherwise there are no limitations on the patients’ underlying conditions.

#### Types of Studies

Parallel-groups RCTs. If a RCT uses a crossover design, latest data from before the first crossover will be used.

#### Types of Interventions

Any antiresorptive pharmacotherapies used to manage bone loss. Treatment arms will include but not limited to: bisphosphonates (e.g. alendronate, etidronate, ibandronate, risedronate, zoledronic acid), denosumab, hormone replacement therapies (HRTs), calcitonin, raloxifene, teriparatide, calcium, vitamin D or D analogs (e.g. calcitriol or alfacalcidol). Because concurrent therapies are common in clinical settings, any combinations of multiple antiresorptive interventions will also be included. Placebo or no antiresorptive therapy will be included as treatment arms as well if data permits. We will not differentiate treatment arms by route or dosage.

### Outcome Measures

#### Change in BMD

We will evaluate change in BMD from baseline, in both percentage and absolute change. BMD change must be calculated based on BMD data collected at the latest follow-up. We will analyze BMD readings taken at the lumbar spine, femoral neck, and total hip. Absolute and percentage change in T-score and Z-score will not be included in this analysis.

#### Fracture Incidence

We will evaluate incidence of vertebral and non-vertebral fracture incidence based on data collected at the latest follow-up. Definitions of vertebral and non-vertebral fractures will be defined as per individual study criteria.

### Search Methods for Identification of Studies

#### Electronic Database Search

We will conduct a librarian-assisted search of Medical Literature Analysis and Retrieval System Online (MEDLINE), Excerpta Medica Database (EMBASE), Web of Science, Cumulative Index to Nursing and Allied Health Literature (CINAHL) and Cochrane Central Register of Controlled Trials (CENTRAL) from inception to September 2019. We will use relevant Medical Subject Headings (MeSH) terms to ensure broad and appropriate inclusions of titles and abstracts (see Supplementary Data).

Major Chinese databases, including Wanfang Data, Wanfang Med Online, China National Knowledge Infrastructure (CNKI), and Chongqing VIP Information (CQVIP) will also be searched using a custom Chinese search strategy (see Supplementary Data).

#### Other Data Sources

We will hand search the reference list of previous meta-analyses and NMAs for included articles. We will also review clinicaltrials.gov and WHO International Clinical Trials Registry Platform (WHO-ICTRP) for registered published or unpublished studies.

### Data Collection and Analysis

#### Study Selection

We will perform title and abstract screening independently and in duplicate using Rayyan QCRI[20]. Studies will only be selected for full-text screening if both reviewers deem the study relevant. Full-text screening will also be conducted in duplicate. We will resolve any conflicts via discussion and consensus or by recruiting a third author for arbitration.

#### Data Collection

We will carry out data collection independently and in duplicate using data extraction sheets developed a priori. We will resolve discrepancies by recruiting a third author to review the data.

#### Risk of Bias

We will assess risk of bias independently and in duplicate using The Cochrane Collaboration’s tool for assessing risk of bias in randomised trials[21]. Two reviewers will assess biases within each article in seven domains: random sequence generation, allocation concealment, blinding of participants and personnel, blinding of outcome assessment, incomplete outcome data, selective reporting, and other sources of bias.

If a majority of domains are considered to be low risk, the study will be assigned a low risk of bias. Similarly, if a majority of domains are considered to be high risk, the study will be assigned a high risk of bias. If more than half of the domains have unclear risk, or if there is a balance of low and high risk domains, the study will be assigned an unclear risk of bias.

### Data Extraction

#### Bibliometric Data

Author, year of publication, trial registration number, digital object identifier (DOI), publication journal, funding sources.

#### Methodology

\# of participating centers, study setting, blinding methods, phase of study, enrollment duration, randomization and allocation methods, technique for BMD measurement, technique for fracture detection.

#### Baseline Data

\# randomized, # analyzed, # lost to follow-up, mean age, sex, # postmenopausal, # taking GC at baseline, mean GC duration at baseline, fracture (vertebral and non-vertebral) prevalence at baseline, mean GC daily dosage at baseline (mg prednisone or equivalent), underlying conditions in the patient population, baseline BMD measurements (in g/cm^2^).

#### Outcomes

Final BMD measurements (in g/cm^2^), percentage/absolute change in BMD from baseline, # with new vertebral fractures at latest follow-up, # with new non-vertebral fractures at latest follow-up.

#### Other Data

Adverse events,description of antiresorptive therapy (i.e.dosage, duration), # taking GC during study, mean GC duration during study, mean GC daily dosage during study (mg prednisone or equivalent), date of latest follow-up.

### Statistical Analysis

#### Network Meta-Analysis

All statistical analyses will be conducted using R 3.5.1[22]. We will perform NMA using the gemtc 0.8-3 library which is based on the Bayesian probability framework[23]. Because we expect significant heterogeneity among studies due to differences in methodology, we will use a random effects model[24]. If data permits, patients receiving no antiresorptive interventions will be used as reference for baseline. If this treatment arm does not exist, placebo patients will be used instead.

We will report BMD results as standardized mean differences (SMDs) with 95% credible interval (CrI) in order to include both percentage and absolute changes in the analysis. This is done to account for studies that fail to report baseline BMD measurements; without baseline BMD measurements, conversion between absolute and percentage changes will not be possible. However, because SMDs are difficult to interpret for most clinicians, we will supplement our BMD results with mean differences (MD) as well, considering only percentage changes in BMD[25, 26]. Fracture incidences will be reported as odds ratios with corresponding 95% CrI. We will run all network models for a minimum of 100,000 iterations to ensure convergence.

#### Network Illustration

We will generate a network diagram for each outcome. A network diagram illustrates treatment arms as variable sized “nodes”, and the trials that provide direct comparisons between arms as “edges” with variable thickness. The size of the nodes represents the total number of direct comparisons that include the represented node, while the thickness of the edges represents the number of publications that offer a direct comparison between the connected nodes.

#### Treatment Ranking

We will use the surface under the cumulative ranking curve (SUCRA) scores to provide an estimation as to the ranking of treatments. SUCRA scores range from 0 to 1, with higher SUCRA scores indicating more efficacious treatment arms[27].

#### Missing Data

We will attempt to contact authors of the original studies to obtain missing or unpublished data. Missing standard deviation values may be imputed using methods described in the Cochrane Handbook for Systematic Reviews of Interventions[28].

#### Heterogeneity Assessment

We will assess statistical heterogeneity within each outcome network using I^2^ statistics and the Cochrane Q test[29]. We will consider an I^2^ index ≥ 50% as an indication for serious heterogeneity, and I^2^ index > 75% as an indication for very serious heterogeneity.

#### Publication Bias

To assess small-study effects within the networks, we will use a comparison-adjusted funnel plot[30]. We will use Egger’s regression test to check for asymmetry within the funnel plot to identify possible publication bias[31].

#### Quality of Evidence

We will use the Confidence in Network Meta-Analysis (CINeMA) web application to evaluate confidence in the findings from our NMA[32]. CINeMA adheres to the GRADE approach for evaluating quality of evidence by assessing network quality based on six criteria: within-study bias, across-study bias, indirectness, imprecision, heterogeneity and incoherence[33, 34]. Although CINeMA utilizes a frequentist approach to NMAs — different from the Bayesian approach used by gemtc — there are no significant differences between frequentist and Bayesian network estimates[35]. We will report the results of our GRADE analysis using a summary of findings table.

#### Meta-Regression

There are several potential factors for increased bone resorption and increased fracture incidences apart from GC therapy, such as gender, post-menopausal status, and age[36]. Prevalent fractures can also increase the risk of subsequent fractures significantly[37, 38]. Variations in these characteristics between studies can result in significant heterogeneity. Therefore, we will conduct meta-regression analyses to check for covariate effects associated with these characteristics.

We will conduct meta-regression on % female in the patient population, % postmenopausal in the patient population, the median age of the population, and the cumulative GC dosage at baseline for both BMD and fracture outcomes. For fracture incidences, we will run a meta-regression on fracture prevalence at baseline. We hypothesize that an increase in mean age, as well as the percentage of females and postmenopausal patients in the population will result in less positive BMD changes and increased fracture incidence. Similarly, an increase in the number of prevalent fractures at baseline will also result in increased fracture incidence.

### Patient and Public Involvement

No patients nor anyone other than the authors and those listed in the acknowledgement section were involved in the design and conduct of this research.

## DISCUSSION

While the latest NMA on this topic have demonstrated that teriparatide and two bisphosphonates (etidronate and risedronate) were effective at reducing fracture incidence and increasing BMD, the authors acknowledged that a lack of data made it difficult to determine the efficacy of treatments such as ibandronate, raloxifene and denosumab[18]. This study aims to significantly expand upon the previous NMA by incorporating all available RCT evidence. While this will not be the first review to evaluate the relative effects of multiple antiresorptive agents among GC users using an NMA approach, it will be the most comprehensive, with new statistical methodologies and multi-language search strategies.

Our review will have several strengths. First, we will include articles of all blinding status and extend our search to major Chinese databases. With their immense patient population and regulations promoting pharmaceutical research, Chinese RCTs are a rich, untapped resource that could greatly strengthen the validity of meta-analyses[39]. Furthermore, we will utilize SMD as a measure of treatment effect for changes in BMD. This will allow us to include both absolute and percentage changes in BMD, even in articles where baseline BMD measurements were not reported. Lastly, we will only include RCT data, and we will use tools such as The Cochrane Collaboration’s tool for assessing risk of bias in randomised trials and CINeMA to evaluate the quality of our included studies and networks.

Our review will also have limitations. Without the restrictions on blinding methods, it is possible that we will include older RCTs that employed open-label or single blinded designs. Because of the potential broad range of publication dates, some of the studies may use outdated techniques and devices to detect fractures and measure BMD, resulting in erroneous readings or significant false positives. Similarly, Chinese RCTs are subjected to the same limitation, as Chinese clinicians may not adopt the same procedures and practices as Western clinicians. Additionally, the patient population in our study will likely have suffered from a variety of different conditions, or may require the administration of drugs other than GCs that can result in GC-unrelated bone loss and fractures.

Despite these limitations, our NMA will be the largest quantitative synthesis assessing antiresorptive therapies among patients undergoing GC therapy to date. It should help physicians and patients with selecting the most effective antiresorptive regimen. Our study may also highlight promising treatments that were not discussed in the previous NMA, providing future researchers with new research directions to further improve clinically relevant outcomes.

## Data Availability

Relevant data are available upon request.

## ETHICS AND DISSEMINATION

The study will not require ethical approval. The findings of the NMA will be disseminated in a peer-reviewed journal and presented at conferences.

## ACKNOWLEDGEMENTS

We would like to offer our special thanks to Alexandra Davis, Library and Learning Centre, The Ottawa Hospital – Civic Campus, for providing her valuable insights on our database search strategy.

## AUTHOR STATEMENT

JWD made significant contributions to conception and design of the work, drafted the work, and substantially reviewed it. EH and ZS made significant contributions to conception of the work, drafted the work, and substantially reviewed it. EZ, KK, AW made contributions to drafting the work, substantially reviewed it, and made revisions to the final work. SS made significant contributions to the methodology of the work. JD, GG made significant contributions to the conception of the work, substantially reviewed it, and made revisions to the final work. All authors read and approved the final manuscript.

## Conceptualization

Jiawen Deng, Emma Huang, Zach Silver, Johanna Dobransky, George Grammatopoulos

## Data Curation

Jiawen Deng, Emma Huang, Zachary Silver, Elena Zheng, Kyra Kavanaugh, Aaron Wen, Stephanie Sanger

## Formal Analysis

Jiawen Deng

## Methodology

Jiawen Deng, Stephanie Sanger

## Project Administration

Jiawen Deng, Johanna Dobransky, George Grammatopoulos

## Supervision

Johanna Dobransky, George Grammatopoulos

## Original Draft

Jiawen Deng, Emma Huang, Zach Silver

## Review & Editing

Jiawen Deng, Emma Huang, Zachary Silver, Elena Zheng, Kyra Kavanaugh, Aaron Wen, Stephanie Sanger, Johanna Dobransky, George Grammatopoulos

